# Longitudinal monitoring exposes correlated temporal protein variations in the female plasma proteome

**DOI:** 10.64898/2026.06.11.26355476

**Authors:** Sofia Kalaidopoulou Nteak, Nicolas Drouin, Stephan Michalik, Henk van den Toorn, Manuela Gesell Salazar, Elke Hammer, Vishnu Mukund Dhople, Silva Holtfreter, Stefan Weiss, Barbara M. Bröker, Grażyna Domańska, Uwe Völker, Albert J. R. Heck

## Abstract

The plasma proteome is a valuable resource for assessment of the physiological state of the donor. Containing hundreds of different proteins of variable concentrations, it displays substantial inter-donor differences in individual protein levels, making each plasma proteome highly donor-specific. Less is known about intra-donor variability in the plasma proteome over time, although such variations may even be more indicative of a changing physiological state. Here we assessed data obtained from the TIMES cohort, comprising 51 apparently healthy participants monitored monthly over 12 months, focusing especially on temporal variations in blood protein levels. Most strikingly, we observed that several women in this cohort revealed strongly correlated temporal variations in their plasma proteome, including most notably PZP, SHBG, FETUB, AGT, SERPINA6, SERPINA7, CP, APOL1 and KNG1, with levels sometimes fluctuating by more than 20-fold. In contrast, such variations were absent in men. Some of the fluctuating proteins have been known to be hormone-regulated (*e.g*., PZP, SHBG), but for others this was not yet fully clear. Through the tight co-variation observed for these proteins in the plasma proteome of women, we can conclude that all these proteins are similarly hormone regulated. The findings reported here not only corroborate previous studies showing estrogen-dependent regulation of several plasma proteins, but also extend this category to include also CP, APOL1, and KNG1. As these latter have been often proposed as candidate biomarkers, they should be validated in sex-balanced cohorts and interpreted with caution, especially in large-scale plasma proteomics studies wherein often only one or a few sampling time points are measured per donor.

## Introduction

Biological and sociocultural differences between the sexes are well established, yet current healthcare systems and research frameworks do often not sufficiently account for these variances.^1,2^ While personalised and precision medicine continue to advance, a substantial number of routine biomarkers are still derived from cohorts lacking a proper sex balance and/or sufficient demographic, ethnic and metabolic variables, potentially limiting their accuracy and clinical soundness. Here, we aimed to address whether and how specific gender biases can potentially also affect blood-based proteome biomarker studies.

Blood circulates through every organ and tissue, acquiring proteins and other molecules along the way. Therefore, blood composition of an individual reflects not only their normal physiology but also particular disease states. Changes in the abundance or modification of specific proteins within the blood can indicate metabolic status, inflammation, organ injury, or even disease progression.^3–5^ Mass-spectrometry based proteomics, but also complementary affinity based proteomics methods, have made it possible to measure in parallel variations in hundreds of proteins, charting the full plasma proteome, which as a whole can be used to monitor and even predict a donor’s (patho)physiological condition, as it reflects genetic predisposition and disease risk.^6,7^ Plasma proteome changes can also be used to monitor the response to therapies and interventions, and to identify for instance diurnal or seasonal rhythms.^3^ Several protein-centric serological assays are already in practical use in the clinic, like for example, C-reactive protein (CRP), which indicates inflammation.^8^

When expanding the application of blood proteome biomarkers, the origins of variation in protein levels must be considered to accurately correlate biomarker behaviour with a given phenotype. An obvious source are genetic differences between donors and several large-scale proteomic studies have tracked how the genome variation impacts the plasma proteome to identify disease-traits that can be used to improve precision medicine.^1^ However, environmental factors play an equally important role in the composition and fluctuations of the plasma proteome. To illustrate, a recent study by Deng et al., showed that age and sex strongly influence protein levels that are also associated with many diseases, so they are biologically important modifiers and potential confounders.^9^ Also, it has been shown that body-mass-index (BMI) and environmental/lifestyle exposures (*e*.*g*. smoking and drinking alcohol) can affect a substantial portion of the proteome, sometimes even more prominently than the genetic background.^10–12^ One other example of such inter-dependence of genetic and environmental factors are exercise and diet, which influence proteins that mediate disease risk, such as type 2 diabetes.^13,14^

Even though nowadays various large-scale plasma proteomics studies do consider different environmental influences, biomarker discovery still faces major challenges. Although advances in sensitivity, throughput and robustness have been made in the last decade, challenges remain, partly caused by the wide dynamic range in protein concentrations and sequence variability between individuals, which hamper accurate and reproducible quantification. Moreover, the field still lacks rigorous study designs, large and longitudinal cohorts, and robust validation models, which leads to few clinically validated markers.^15,16^ These challenges cause a considerable translational bottleneck. For example, they limit our knowledge about potential female-specific variation in the plasma proteome, which might result in a bias towards the male proteome as a base for biomarker discovery.

The focus of the present work is to identify potential differences and/or longitudinal fluctuations in the blood proteome related to the sex of the donor. It is already known that hormonal contraceptives can impact certain blood protein concentrations, notably those of the sex hormone-binding globulin (SHBG) protein and the pregnancy zone protein (PZP).^17,18^ Sex-specific differences in the blood proteome are typically due to either tissue-specific proteins or to those involved in hormone-related pathways. Some of the most notable female-male differences arise from proteins that are abundantly expressed in tissues unique to one sex, like prostate-specific antigen (PSA) in males and follicle-stimulating hormone (FSH) in females. However, also other hormone- and metabolism-linked proteins such as leptin (LEP) and adiponectin (ADIPOQ) have been reported to show consistent differences when considering sex.^19^ Additionally, the female proteome can be enriched with hormone-regulated proteins, that are influenced by menstrual cycle, pregnancy, or by hormonal therapies. During pregnancy, the abundance of several maternal plasma proteins shifts with gestational age, reflecting changes in inflammatory tone, immune response, vascular biology, and hormone-related processes.^20^

To study the effects of sex on the plasma proteome of healthy adults, we applied an unbiased and sensitive technique to the longitudinal TIMES cohort.^21^ In more detail, we used a semi-automated sample preparation workflow together with a robust and high-throughput LC-MS/MS set up to monitor the proteome of 51 adults monthly over a period of a year. All samples were obtained in 2017/2018, *i*.*e*., prior to the COVID outbreak. Cumulatively, this set of around 600 samples provided a longitudinal view of the personalized plasma proteome of 51 donors. We confidently identified several sex-related inter- and intra-donor variations, of which some have been previously linked to hormonal regulations, while others had so far not yet been clearly recognised as such. We find that in some female donors these proteins display temporarily co-regulated elevated levels over the full year. Nearly all our findings could be validated in an independent longitudinal cohort, the AICOVI cohort that also contained solely healthy donors. Our study emphasizes the need to take into account temporal sex-related differences to enhance the quality of clinically relevant biomarkers in personalised medicine to avoid gender bias.

## Experimental Procedures

### Study design

In the TIMES study (Tracking Individuals Monthly for Evaluating Stability), whole blood samples were obtained from 51 adult volunteers at the University Medicine Greifswald (19 males and 32 females) aged 19-56 years. The longitudinal samples were collected over a period of one year with 12 consecutive monthly samplings from June 2017 through May 2018. In total 587 EDTA blood samples could be collected and investigated by plasma proteomics. Detailed characteristics of the study cohort have been described previously.^21^ For validation of the findings, a second cohort was studied, consisting of 11 health care workers that were part of the AICOVI study (Adaptive Immune Response after COVID-19 Vaccination), which was also conducted at University Medicine Greifswald. The AICOVI participants (4 males and 7 females) volunteered to donate plasma longitudinally over a period of a year in 2021, at 9 different time points. These volunteers all received three doses of a COVID-19 vaccine. They remained healthy—as far as known—during the whole study period. In total 99 blood samples from the AICOVI cohort were prepared and analyzed^22,23^ as for the TIMES cohort proteomics study. The AICOVI cohort primarily served to cross-validate findings we extracted from the TIMES cohort study.

### Plasma sample preparation

Plasma samples were processed using a previously described high-throughput proteomics protocol.^21,24^ Briefly, 10 µL of plasma was diluted 1:1 with Tris buffer (100 mM, pH 8.5). 4 μL of the diluted sample was combined with lysis buffer containing 4% sodium deoxycholate (SDC), 160 mM chloroacetamide (CAA), 100 mM Tris, pH 8.5, and 40 mM tris(2-carboxyethyl)phosphine (TCEP). The samples were heated to 95 °C for 10 minutes to denature the proteins and to allow reduction and alkylation to occur. Following this step, the samples were further diluted with Tris buffer. Trypsin (Sigma-Aldrich) and LysC (Wako) were added to each sample at a final amount of approximately 2 µg total protease (10 µL of a 0.2 µg/µL enzyme solution). Protein digestion was carried out overnight at 37 °C and was terminated by the addition of trifluoroacetic acid. The resulting peptide mixtures were then diluted 36-fold in 1% TFA prior to downstream analysis. The analysis was performed in five batches (two 96-well plates/batch). To monitor batch effects we included six QC samples of pooled serum (HSER-CUSTOM, amsbio) in each well plate.

### LC-MS/MS DIA for proteomics

Approximately 200 ng of peptides in a total volume of 20 µL were loaded onto Evosep® Pure tips following the manufacturer’s instructions. Samples were analyzed using an Evosep One liquid chromatography system (Evosep, Odense, Denmark) coupled to a timsTOF HT mass spectrometer (Bruker Daltonics, Bremen, Germany). Data were acquired in dia-PASEF mode. After elution from the Evosep® Pure tips, peptides were separated on an EV-1109 analytical column (ReproSil Saphir C18, 1.5 µm particle size, 8 cm × 150 µm inner diameter) using the Evosep 60 SPD method. Chromatographic separation was performed with mobile phase A consisting of 0.1% formic acid in water and mobile phase B consisting of 0.1% formic acid in acetonitrile. Peptides were ionized using a captive spray source operated at 1,600 V, with a drying gas flow of 3 L/min and a source temperature of 180 °C. The dia-PASEF acquisition scheme and ion mobility settings were defined as detailed in Supplementary Table 3. Precursors were selected across an m/z range of 300–1200 using 12 dia-PASEF scans, corresponding to an 8.3% duty cycle. The ion mobility range was set between 0.6 and 1.6 Vs/cm^2^, with both accumulation and ramp times set to 100 ms. Collision energy was applied using the default mobility-dependent ramp (20–59 eV). Data acquisition was performed with synchronized accumulation and ramp times of 100 ms, resulting in a 100% duty cycle.

### Data processing and statistical analysis

Raw data analysis was done using DIA-NN (version 1.9) in library-free mode.^25^ Trypsin/P was used as a protease and up to two missed cleavages were allowed. Cysteine carbamidomethylation and methionine oxidation were used as fixed and variable modifications, respectively. The mass accuracy was set to 20 ppm and the MS1 to 10 ppm. The unrelated runs, isotopologues, match-between-runs and no shared spectra options were enabled, whereas the heuristic inference was disabled. The protein inference was set to Genes, and the IDs, RT and IM profiling was used for the library generation. The FDR was at the default 1%. The data was searched against an in-house prepared blood protein database.^21^ Briefly, this database consists of SwissProt sequences of the 2,445 most common blood proteins according to the Human Protein Atlas (acquired November, 2023), and their 5% most frequent single amino-acid variations found in the NextProt database^26^ (acquired May 2024). In addition, immunoglobulin sequences covering the more constant regions were manually added into this database. Protein sequences describing just the variable domains of immunoglobulins were omitted. The main report of DIA-NN was filtered from the contaminants and from proteins with less than 2 identified peptides and used for further analysis. The abundance of each protein was calculated based on MaxLFQ values using the diann_maxlfq function of the DIAgui package (https://github.com/mgerault/DIAgui), using Q.Value, Lib.Q.Value and Lib.PG.Q.Value, all set at 1% to filter precursors. The log_10_-scaled MaxLFQ values of each protein were then converted into absolute concentrations through a linear model, using the average protein concentrations of 22 known and robustly quantified plasma proteins by Gaither et al.^27^ Statistical analyses were performed in R using the packages dplyr v1.1.4, rstatix v0.7.3, and FSA v0.10.0. Protein concentrations were compared between groups using a non-parametric approach. Specifically, differences between sexes were assessed for each protein using the Kruskal-Wallis test and pairwise comparisons were conducted using Dunn’s test. To account for multiple testing across all proteins, p-values from the pairwise comparisons were adjusted using the Benjamini-Hochberg procedure to control the false discovery rate. For effect size interpretation, fold changes between female and male groups were calculated based on mean protein concentrations. For variability assessment between donors, the inter-donor coefficient of variation (CV) was calculated for each protein and sex group as the standard deviation to the mean of donor-averaged protein concentrations. Data visualization was performed via the ggstatsplot v0.13.4 and ggplot2 v4.0.3 R-packages.

## Results

### Study design of the TIMES cohort

Here we set out to investigate potential dynamic changes in the plasma proteome of apparently healthy female and male individuals of the TIMES cohort (Tracking Individuals Monthly for Evaluating Stability) using a previously published workflow.^21^ This TIMES cohort consisted of 51 individuals, namely 32 females and 19 males, from whom monthly blood samples were collected over one year. The participants had an average age of 34.0 years and no known disease or medication treatment. Inclusion criteria, as previously described,^21^ included (i) age 18–60 years, (ii) written informed consent, and (iii) no acute illness in the last seven days before sampling. Exclusion criteria included (i) body mass index <18.5⍰kg/m^2^, and (ii) presence of blood coagulation disorders, anemia or similar conditions. Plasma samples were obtained from a total of 53 participants once per month for 1 year (June 2017–May 2018). The plasma proteome profiles of all individuals at all time points (∼600 samples), together with the 58 quality control samples were prepared on an automated liquid handling system and analysed with liquid chromatography-MS analysis using equal plasma volumes and a DIA-based strategy. As we were interested mostly in the analysis of common plasma proteins,^28^ the acquired proteomics datasets were searched against an in-house generated targeted blood protein database containing 2,445 entries.^21^ After stringent filtering, 224 genuine plasma proteins could be confidently identified and robustly quantified at all sampling points of the TIMES cohort. An overview of this dataset has been published earlier.^21^ Here we report the results of a novel, sex-specific analysis.

### Sex-related differences in the plasma proteome

Taking advantage of the features of the longitudinal TIMES cohort, we aimed to elucidate sex-related differences in the plasma proteome analysing the data at three different layers. First, we assessed the overall differences in protein levels between sexes. These were assessed by grouping all samples (independent of donor and time point) according to male or female sex of the donor and determining differences in mean protein concentrations between the two groups. Then we examined the inter-donor variability in both male and female subgroups, and lastly the intra-donor variability over the time-course of one year.

### Global differences in protein levels between sexes

To uncover proteins that exhibit significant overall differences in plasma concentrations between females and males, we identified proteins that had statistically significant higher concentrations in the female donors (Figure 1). This analysis led to a list of 9 proteins including, as expected, the well-known pregnancy/oestrogen-related proteins PZP, and SHBG. Also, the concentrations of fetuin-B (FETUB) and angiotensinogen (AGT) were found to be higher in women, in line with more recently reported data.^17^ Next to these previously described proteins, corticosteroid-binding globulin (SERPINA6), thyroxine-binding globulin (SERPINA7), ceruloplasmin (CP), apolipoprotein L1 (APOL1) and kininogen-1 (KNG1) exhibited significantly higher concentrations in the female than the male donors with p-values below 0.001 (except APOL1 where p ∼0.05). For reference, we depict in Supplementary Figure 1 the data for the abundant plasma proteins immunoglobulin heavy constant gamma 1 (IGHG1), haptoglobin (HP) and apolipoprotein A-II (APOA2), whose concentrations did not differ between the sexes in the TIMES cohort. Of note, the data presented in Figure 1 represent protein concentrations of all 364 female and 221 male plasma samples, respectively; including all time points. Strikingly, the variation in plasma protein concentrations was much higher in females than in males. The fold-changes between the means of the two groups are also shown in Figure 1. They illustrate that the low abundance of PZP and SHBG in the male donors leads to the largest fold changes of about 59-fold and 5-fold increase in women, respectively. The fold changes of the other 7 proteins are lower, between 1.1 and 2.6, but still significant (Figure 1).

**Figure 1.**
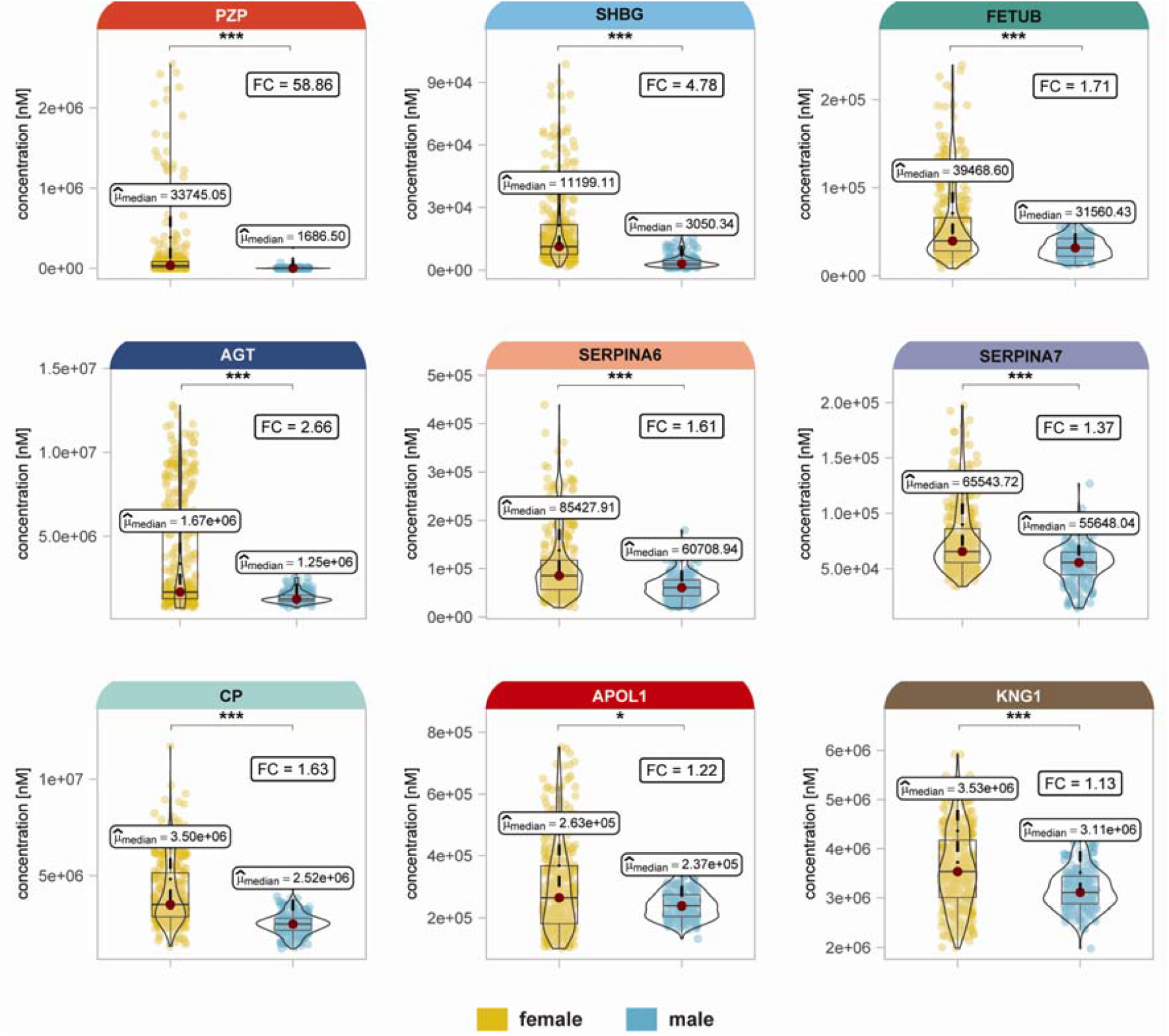
Differences in overall female and male plasma protein concentrations. Distribution and median concentrations of proteins found to be significantly more abundant in the plasma proteome of women compared to men. The median concentrations for each protein and sex is highlighted, and its value is displayed 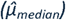. One star (*) represents p-value (Benjamini-Hochberg (BH) adjusted) below 0.05, two stars (**) below 0.01, and three stars (***) below 0.001. The most distinctive sex-related proteins include PZP, SHBG, FETUB, AGT, SERPINA6, SERPINA7, CP, APOL1 and KNG1 all exhibiting p-values below 0.001, except of APOL1 with a p-value below 0.05. The fold change (FC) between women and men is annotated in the top-right box. For comparison, data for three other plasma proteins (IGHG1, HP and APOA2), which show no differences in median plasma protein concentrations between female and male donors, are provided in Supplementary Figure 1.

For validation we examined 11 donors of an independent cohort (AICOVI) and calculated the p-values for the same 9 proteins, in this case sampled at 9 time points over a period of one year (99 plasma proteomes, Supplementary Figure 2). Also, in this smaller cohort the plasma concentrations of PZP, SHBG, AGT and CP were higher in women than in men with p-values of less than 0.001, whereas SERPINA7 had a p-value of less than 0.05. In the small AICOVI cohort FETUB, SERPINA6, APOL1 and KNG1 did not differ significantly between the sexes (likely due to limited sample size and a sex-imbalance), but the median values in the female group were still higher than those in the male individuals (Supplementary Figure 2).

### Inter-donor variability in the male and female subgroups

Figure 1, showing the samples of all donors and all time points, revealed a much wider spread in protein concentrations of female than male donors. To better understand this, we plotted the mean concentrations of each donor separately (Figure 2), with the data for females in yellow and those for males in blue. Clearly, the inter-donor variability is very low in males, while in females the concentrations of these 9 proteins show pronounced inter-donor variability. Illustratively, PZP showed the largest coefficient of variation between female donors (mean CV: 227%), with donors 33 and 49 having a ∼10-fold higher concentration than the female average. Other female donors exhibited average plasma concentrations for these 9 proteins on par with those measured in the male samples (*e*.*g*., female donors 5, 53 and 56). Similarly, in the AICOVI cohort all male donors have a low inter-donor CV for these 9 proteins, whereas the female donors showed substantially more variation (Supplementary Figure 3).

**Figure 2.**
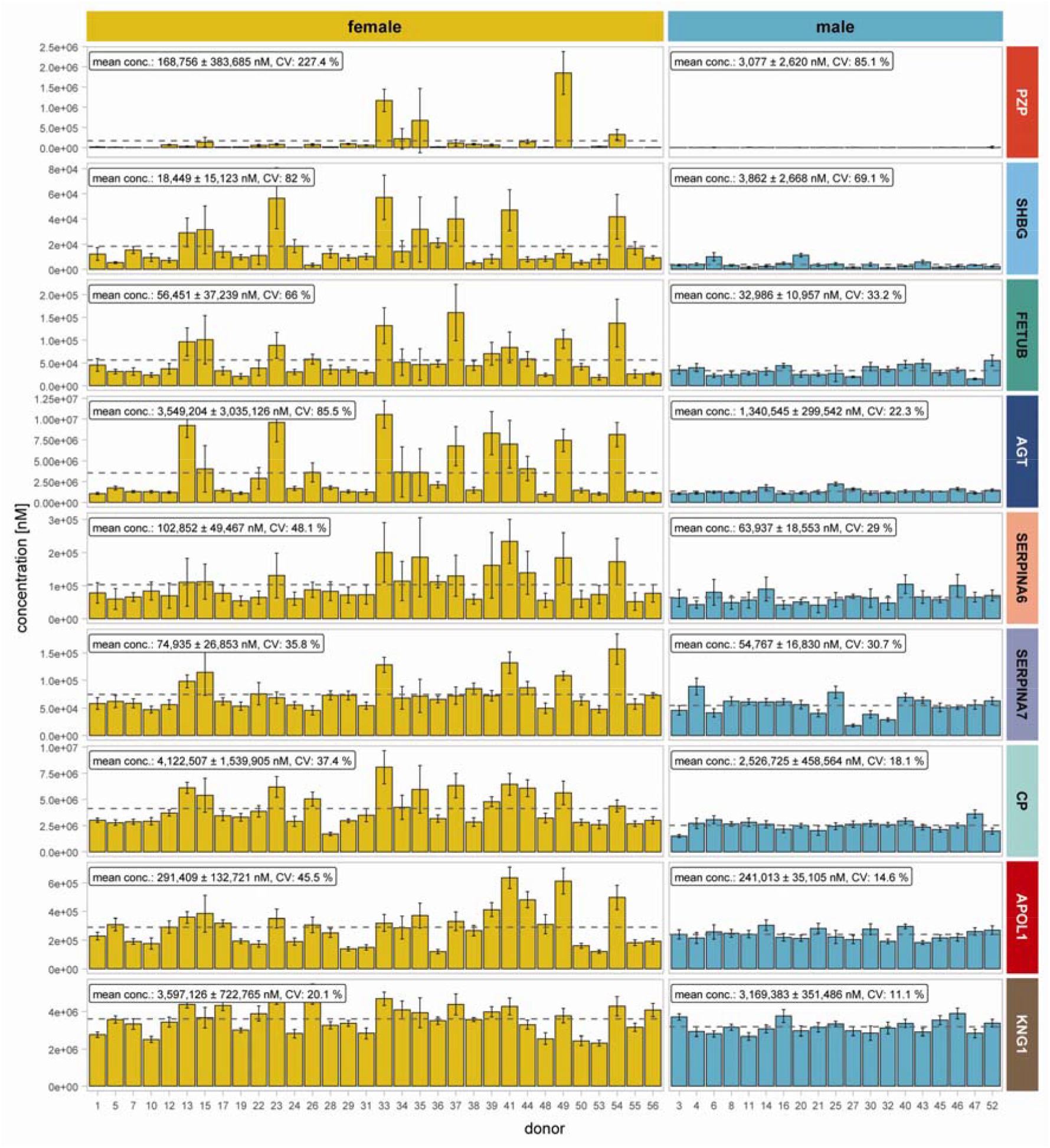
Inter-donor variability in female and male plasma protein concentrations. One-year average plasma concentrations and standard deviation (error-bars) of the 9 sex-affected proteins here depicted per individual female (yellow) and male (blue) donors. The dashed line signifies the overall mean concentrations across all females and males. At the top the overall mean concentration is given, together with the observed variation which is substantially higher when analysing data from female donors. Strikingly, a sub-group of female donors seems to be responsible for the elevated levels of all these 9 proteins, well above the median of all female donors (e.g., donor 33 and 49 and 54), whereas other female donors exhibit concentrations alike those observed in the male donors.

We conclude that in some, but not all, women the mean plasma concentrations of these 9 proteins are substantially higher than in men. This explains the significant differences between the sexes that are shown in Figure 1.

### Intra-donor high longitudinal variability occurs exclusively in a female subgroup

We next focused on these apparently aberrant female donors and monitored the temporal behaviour of the same 9 plasma proteins during the year of monthly sampling. In Figure 3 (and the upper panel of Supplementary Figure 4) we show 5 representative female and 4 male donors. The female donors were of three categories: *a*. exhibited high intra-donor variance (female donors 34, and 35), *b*. showed low variance but high concentrations of these 9 proteins (female donor 54), and in *c*. both intra-donor variance and protein concentrations were low and similar to the values observed in men (female donors 7, and 29). For reference in Figure 3 we also included 4 male donors that were randomly selected, as all male donors exhibited low temporal intra-donor variability protein profiles for these proteins (male donors 3, 30, 32, and 47; Supplementary Figure 4, bottom panel). The data depicted in Figure 3 reveal that the longitudinal profiles of the 9 proteins are correlated in the selected female donors. Donors 34 and 35 exhibit contrasting seasonal profiles. Donor 34 shows elevated protein levels between June and October, while donor 35 has an increased protein concentration between January and May. Since the changes are occurring randomly across the year, we assume that there is no seasonal regulation of these proteins. In contrast, the female donors 7 and 29 display stable profiles over time, similar to the male donors. Interestingly female donor 54, shows also a stable longitudinal profile, but with markedly elevated baseline protein concentrations. These observations are highlighted in Figure 3 in a subset of donors, but similar patterns are observed in the complete TIMES cohort (Supplementary Figure 4, lower panel).

**Figure 3.**
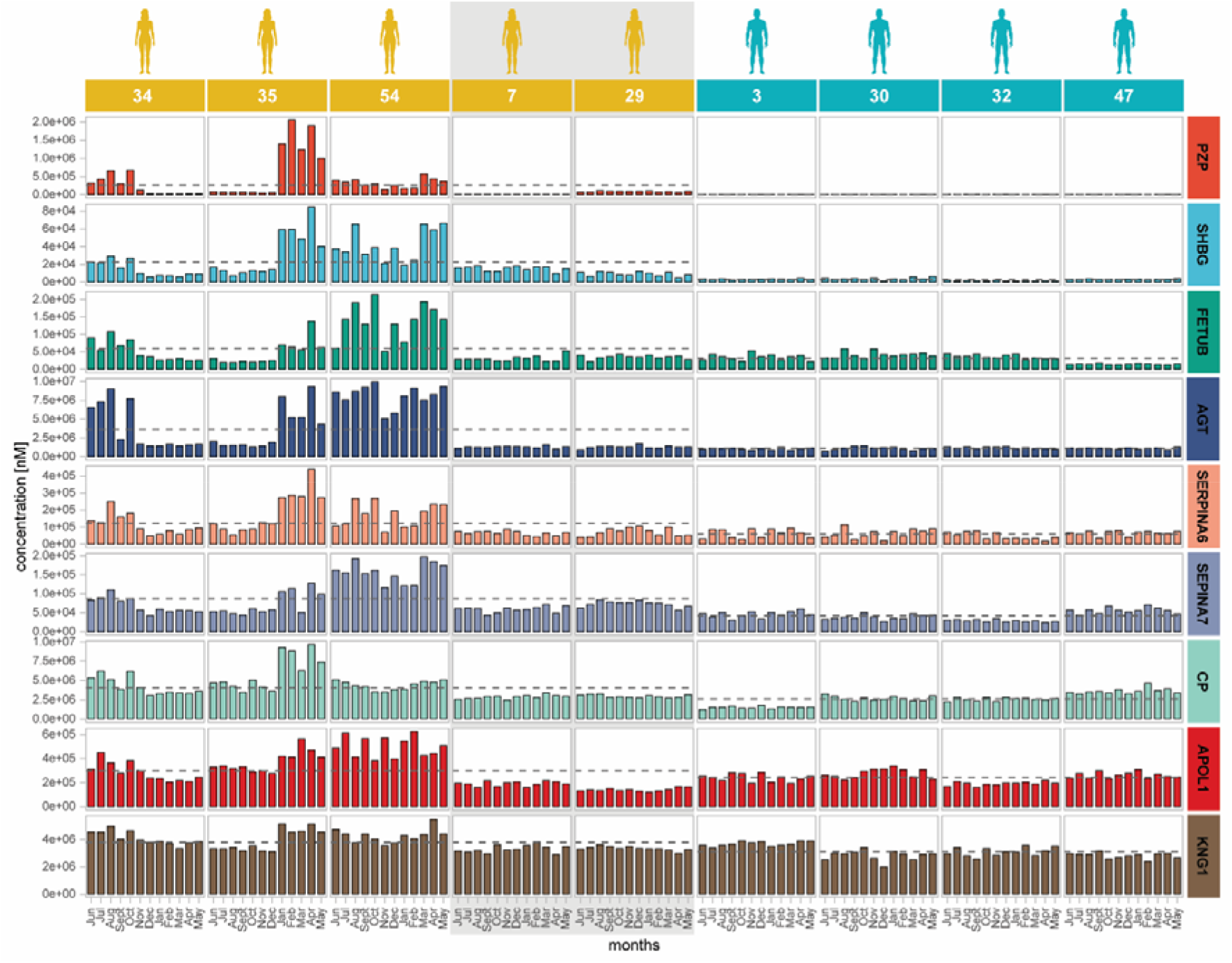
Tightly correlated temporal fluctuations in a set of plasma proteins observed in selected female donors. Bar plots of plasma protein concentrations in selected donors reveal temporal bursts and declines in some female donors. Illustrative data from 5 female donors and 4 male donors are shown. The first three female donors were selected as they showed high temporal variance, the last two as they showed very little variance. The male donors were randomly selected, since all males show similar protein patterns. Each graph represents the concentrations at each of the 12 time points of sampling.. In some female donors (e.g., donor 34, 35) sudden bursts in protein abundance occur, that are not synchronized between the individuals. However, in each female individual, the bursts in protein abundance correlate well between the 9 proteins.

To highlight this tight co-regulation, we plotted the concentrations of these proteins in all donors against the respective AGT concentrations as a reference (Figure 4). AGT was chosen as a reference protein, since it has been previously shown to be higher in female donors and it has the highest overall abundance among the 9 selected proteins. The concentrations of all other 8 proteins were highly correlated with those of AGT in the female donors, albeit mostly in the subset of data where the plasma levels are elevated. FETUB and CP show the highest correlations with plasma AGT concentrations for the female donors (r_s_= ∼0.75), while SHBG, SERPINA6-7, APOL1 and KNG1 have r_s_ values in the range of 0.5-0.6. PZP exhibits a more complicated pattern, since overall, it does not correlate with AGT, but in certain female individuals like donors 34, 35, 38, 44, 22, 15 and 17 the correlation is more than 0.7 across the time points. In the AICOVI validation cohort (Supplementary Figure 6) the coefficients of correlation in the female donors are similar to the TIMES.

**Figure 4.**
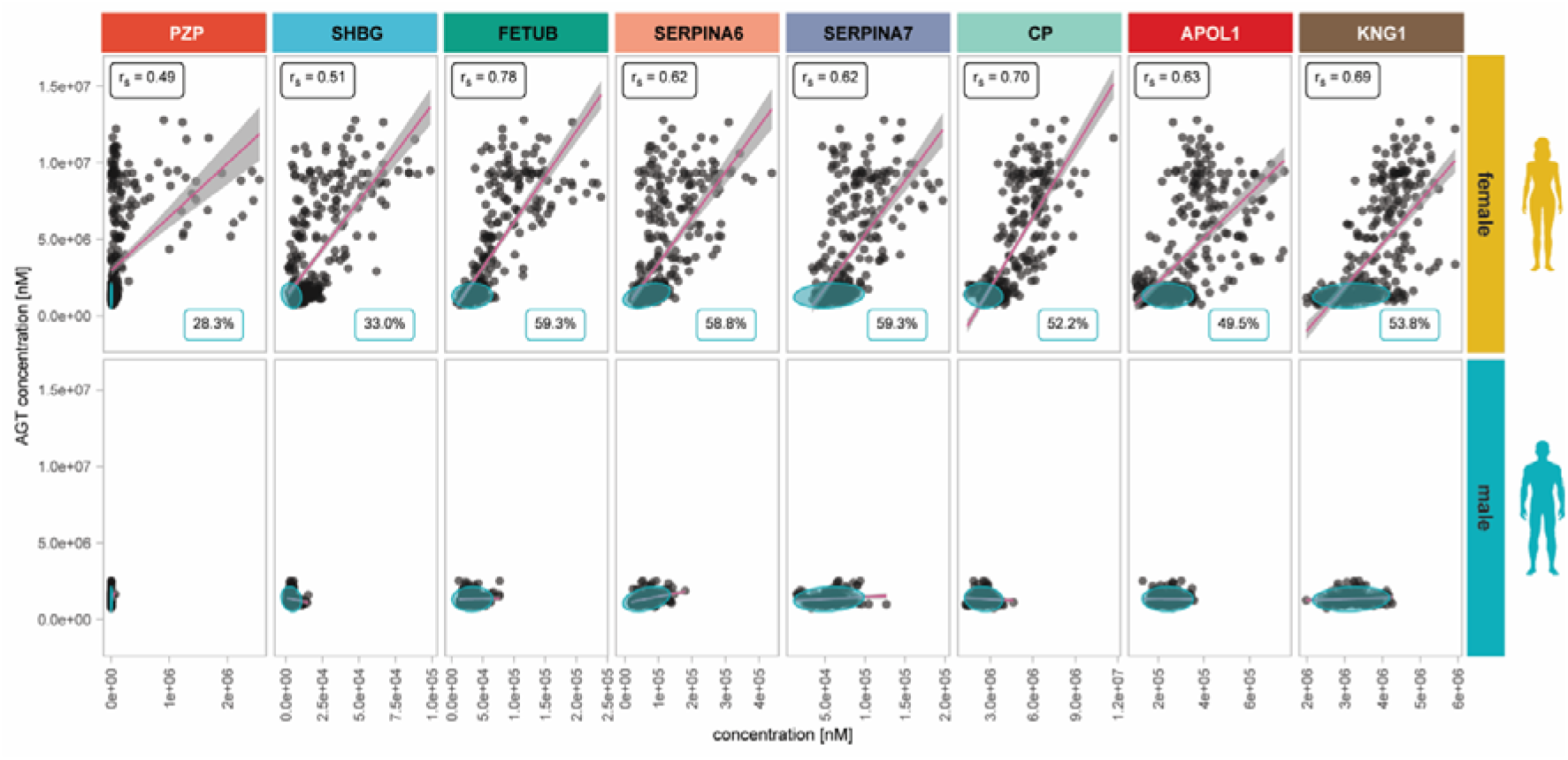
Correlation plots of eight selected plasma proteins with AGT in female and male individual donor samples. The plasma concentrations of 8 plasma proteins are plotted against the corresponding AGT concentrations at each sampling. In female donor samples (upper panel) the observed correlations are high. In the male donor samples (bottom panel) there is very little variation, and thus low correlation. The blue ellipses depict the 95% confidence region of the concentration ranges in males, calculated assuming a bivariate normal distribution. The percentages at the bottom right indicate the fraction of female donor samples that fall into the boundaries of the “normal male” distribution. Spearman correlation coefficients (rounded to two decimal places) are shown as r_s_.

As not all female donors showed a uniform regulation of these 9 proteins, we next examined all male and female AGT-protein relationships using a covariance-based approach. For each of the 8 proteins, we fitted a 95% confidence ellipse to the AGT-protein distribution in the male donors, assuming a bivariate normal distribution. These ellipses estimate the ranges of protein covariance observed in males. We next calculated the proportion of female donor samples whose AGT protein measurements fall within the ellipse boundaries of the male donors. This percentage reflects the fraction of female donor samples whose AGT-protein relationship corresponds to the “normal male” variation (ellipse). This approach highlighted that the plasma levels of PZP, SHBG, FETUB, SERPINA6, SERPINA7, CP, APOL1, and KNG1 vary only little between the male samples (Figure 4, bottom panel). Around half of the female samples (30-60% for individual proteins) show a similar behaviour. Thus, the overall higher levels of these proteins in female samples shown in Figure 1 are due to elevated levels in a ∼50% subset of the female samples. In these female samples all 9 proteins are elevated concurrently.

The tight correlation in the dynamic fluctuation of the 9 plasma proteins likely means that their synthesis is controlled by alike stimuli. All these proteins are synthesized in the liver, before they are secreted into the circulatory system.^29^ As it is well established that the expression of PZP and SBHG is strongly regulated by oestrogen, our data strongly suggests that the synthesis and secretion of the other 7 proteins is similarly controlled.^30,31^ In the TIMES study we did not monitor menstrual cycle, pregnancy, menopause or use of contraceptives, nor did we measure the levels of sexual hormones. Therefore, while sexual hormones could be a decisive factor, this hypothesis remains to be tested in future studies.

Most observations made in the TIMES cohort were reproduced in the AICOVI participants, including the tight temporal co-variation in PZP, SHBG, FETUB, SERPINA6, CP, and KNG1 with AGT in the female donors (Supplementary Figures 5 and 6).

## Discussion

In this study we used plasma proteomics to expose potential temporal differences in protein levels, focusing on observed sex-related differences. We used data obtained from the TIMES cohort, comprising of 51 supposedly healthy participants sampled monthly over one year. In an earlier study of the same cohort, we observed that in these healthy adults the concentrations of most plasma proteins were stable over time for the timespan of one year.^21^ The individual stability was in sharp contrast with a large inter-donor variability of the same plasma proteins. The large family of immunoglobulins (*e*.*g*., IgG, IgA, and IgM, and their sub-class variants) are good examples for this.^21^ The two hallmarks of the plasma proteome define each donor’s personal plasma proteome like a finger print as revealed by unsupervised clustering. This analysis showed that temporal sampling points clustered mostly very tightly together, whereas different donors separated greatly from each other. Thus, generally the inter-donor variability is high, intra-donor longitudinal variability is low. We thus concluded that the most appropriate reference sample for a diseased individual is a sample obtained from the same donor at an earlier healthy state. While previous studies have hinted at similar results,^32–34^ the TIMES cohort is unique in including a large number of longitudinal sampling time points from a cohort of apparently healthy donors. Comprising 19 male and 32 female participants, the TIMES has no strong sex bias, which is not always the case in long-term proteomics studies.^35^

We hypothesized that these distinctive features of the TIMES cohort would allow us reveal differences in the plasma proteome between men and women. As an initial step, we compared average plasma protein concentrations between men and women and identified 9 proteins, namely, PZP, SHBG, FETUB, AGT, SERPINA6, SERPINA7, CP, APOL1, and KNG1, that were present at higher levels in women than in men (Figure 1). Upon closer examination it became apparent that these sex differences were largely driven by a subset of female donors (Figures 2-4). Moreover, the elevated protein concentrations were not stable but fluctuated highly over time in some women, in some cases there were more than 20fold changes over the course of one year. Because of the longitudinal design of the study with 12 consecutive monthly samplings, we were able to examine temporal relationships between these proteins. Interestingly, the dynamic changes in the concentrations of the 9 proteins were strongly correlated with one another, indicating a shared regulatory influence rather than independent regulation.

It is well established that PZP, SHBG, and SERPINA6 are strongly influenced by sex hormone physiology. PZP and SHBG have long been recognized as oestrogen-responsive proteins. They show markedly higher concentrations in women than in men and increase substantially in response to oestrogen exposure, such as oral contraceptive use or pregnancy.^20,36^ In healthy men, PZP circulates at baseline concentrations far below the levels observed during pregnancy or high oestrogen states.^37^ Similarly, SERPINA6 (also known as transcortin) plays a key role in the transport of glucocorticoids and exhibits sex differences in expression and physiological effects.^38^ Beyond these canonical sex hormone-regulated proteins, other plasma proteins such as FETUB and AGT have also been linked to hormone status. Recent large-scale proteomics studies have shown that levels of both FETUB and AGT are increased in women using hormonal contraceptives, indicating their responsiveness to hormonal modulation.^17^ In the present study, we extend this list of putatively hormone-associated proteins to include CP, APOL1, KNG1, and SERPINA7, which exhibited similar inter- and intra-individual sex-dependent patterns. Additionally, we observed strong correlations among certain plasma proteins, most prominently between FETUB and AGT, highlighting coordinated regulation that may reflect shared hormonal or physiological influences.

Somewhat less known, CP, APOL1, KNG1 and SERPINA7 display a similar pattern to the well-known oestrogen-related proteins, and they might be therefore also involved in hormonal regulation. Of these, CP (ceruloplasmin) is the primary carrier of copper in the bloodstream. It is often used as a biomarker for various copper-related disorders but also for certain inflammatory conditions and other diseases.^39^ It has been described that pregnancy may lead to elevated levels of CP as well as oestrogen or contraceptive therapy.^40^ However, elevated CP levels are also discussed as biomarkers for leukaemia and Hodgkin lymphoma.^41,42^ Here we show that if CP is used as biomarker for diseases, the sex of the patients must be carefully considered as well as physiological fluctuations in some women. In agreement with other studies,^43^ we find in both the TIMES and AICOVI cohorts a clear higher concentration and inter donor variance of CP in female donors (TIMES; female median conc.: 4,122,507 nM; mean CV: 37.4%, AICOVI; female mean conc.: 3,503,013 nM; mean CV: 36.3%) than males (TIMES; male mean conc.: 2,526,725 nM; mean CV: 18.1%, AICOVI; male mean conc.: 1,943,783 nM; mean CV: 4).

Also, KNG1 and APOL1 have been reported as potential biomarkers in several contexts, but they are not yet routinely established clinical biomarkers. KNG1 has for instance been associated with advanced colorectal adenoma and early colorectal cancer (CRC), with altered pre-versus postoperative levels in CRC patients.^44^ APOL1 (both gene variants and protein levels) is an increasingly supported biomarker in specific settings (*e*.*g*., renal genetics, thyroid cancer prognosis, experimental liver fibrosis).^45^ We propose that also for these proteins the sex of the patients must be carefully considered, as well as temporal variations in the levels of these proteins in the plasma of some female donors.

Notably, although these 9 proteins exhibit coordinated abundance patterns in plasma, their genes do not share single conserved regulatory sequences. Moreover, these genes are located on different chromosomes and are not arranged as a single tight genomic cluster. Instead, their co-regulation likely reflects the influence of common hepatic transcription factors and hormonal or inflammatory signalling pathways. In particular, thyroid hormone receptors, oestrogen signalling, and/or cytokine-mediated regulation likely lead to correlated plasma levels.^46,47^

## Conclusions

In this study, we used the longitudinal design of the TIMES cohort to investigate intra-donor sex-dependent variability in the plasma proteome of healthy adults. While inter-donor differences are a well-recognized feature of plasma proteomics, our results demonstrate that temporal variations within individuals can provide equally important biological insight. Using monthly sampling over one year, we identified a distinct set of plasma proteins (*e*.*g*., PZP, SHBG, FETUB, AGT, SERPINA6, SERPINA7, CP, APOL1, and KNG1 that exhibited pronounced and tightly correlated temporal fluctuations in a subset of female donors, but not in male individuals. These dynamic changes were substantial, with protein concentrations varying by more than 20-fold over time.

Several of the identified proteins are known to be hormone-regulated, particularly by oestrogen, and we confirmed their higher average abundance in females in an independent cohort. Importantly, our longitudinal data revealed that these proteins are not uniformly elevated in all women; instead, increased levels occurred only in some females and at discrete time points. The strong co-variation among these proteins suggests shared regulatory mechanisms, most likely driven by hormonal influences. Our study therefore extends previous reports to include CP, APOL1, and KNG1 in the group of proteins regulated by female hormones.

These findings have important implications for plasma proteomics and biomarker discovery. Hormone-driven temporal variability may confound ross-sectional analyses and can thus lead to misinterpretation of physiological fluctuations as disease-related signals. Incorporating sex and hormonal context into study design and data interpretation will be essential for improving the robustness and clinical relevance of plasma proteomics studies.

## Supporting information

All supplemental Figures

## Data Availability

All proteomics data were previously deposited in an accompanying study on the sane TIMES cohort, notably in this article
Michalik, S., Kalaidopoulou Nteak, S., Drouin, N. et al. Immunoglobulin sub-class levels define inter-donor plasma variability: a longitudinal dual-lab study. Mol Syst Biol (2026). https://doi.org/10.1038/s44320-026-00218-5. As stated in the current manuscript under the data availability paragraph: Restrictions apply to the availability of data generated or analysed during this study to preserve patient confidentiality or because they were used under license. Access to the data is permitted under controlled access in accordance with the European Union General Data Protection Regulation (GDPR) at https://ship.limequery.org/532462.

## Limitations of this study

Despite the robust longitudinal design and comprehensive proteomic profiling, our study has limitations. First, the cohort size, while adequate to detect intra- and inter-individual variability, is limited and may constrain the generalizability of the findings. In addition, the cohort represents a restricted range of demographic and health characteristics, and detailed information on factors such as menstrual cycle, pregnancy, menopause or hormonal contraceptive use, or other sex-specific physiological conditions was not accessible because the study was designed to protect the participants’ privacy. Furthermore, we focused in our studies primarily on the 200-300 most abundant plasma proteins to ensure robustness of the measurements and to avoid “missing values”. Nowadays, deeper plasma proteomics can be performed, which is expected to uncover additional genes/proteins that exhibit sex-related differences and temporal fluctuations in the plasma proteome.

## Data availability

Restrictions apply to the availability of data generated or analysed during this study to preserve patient confidentiality or because they were used under license. Access to the data is permitted under controlled access in accordance with the European Union General Data Protection Regulation (GDPR) at https://ship.limequery.org/532462.

## Supplementary data

The supplementary information contains the Supplementary Figures 1-6.

## Acknowledgements

This research received funding through the Dutch Research Council (NWO) through funding the Roadmap programs X-omics (project 184.034.019) and BioBeyond_NL (grantnumber 184.037.015). AJRH acknowledges additional support from NWO through the Spinoza Award SPI.2017.028. We thank Celine Miedema for critically reviewing and editing the manuscript.

## Author contributions

SKN, SM, UV, AJRH conceptualization and writing final draft;

SKN, AJRH writing – original draft;

SKN, SM formal analysis and data visualization;

SH, GD study design and acquisition of participants’ data and biomaterials

SKN, SM, ND, HT, MGS, EH, VD, SH, SW, BMB, GD, UV sample preparation and reviewing final draft; SKN, SM, ND, HT data curation;

AJRH, UV, BMB resources;

