## Supplementary material for "Longitudinal monitoring exposes correlated temporal protein variations in the female plasma proteome": All supplemental Figures

### Table of Contents


| | Page |
|---|---|
| **Supplementary Figure 1 \|** Plasma protein levels of three proteins (IGHG1, HP and APOA2) that do not show any differences in median protein concentrations between female and male donors. | **S3** |
| **Supplementary Figure 2 \|** Differences between female and male plasma protein concentrations in AICOVI cohort. | **S4** |
| **Supplementary Figure 3 \|** Averaged plasma concentrations of selected proteins per female and male donors in AICOVI cohort. | **S5** |
| **Supplementary Figure 4 \|** Radar plots of selected proteins in all (panel below) and selected (panel above) donors. | **S6** |
| **Supplementary Figure 5 \|** Bar plots of selected plasma protein concentrations in all individuals of AICOVI cohort. | **S7** |
| **Supplementary Figure 6 \|** Correlation plots of selected plasma proteins with AGT in female and male individuals of the AICOVI cohort. | **S8** |

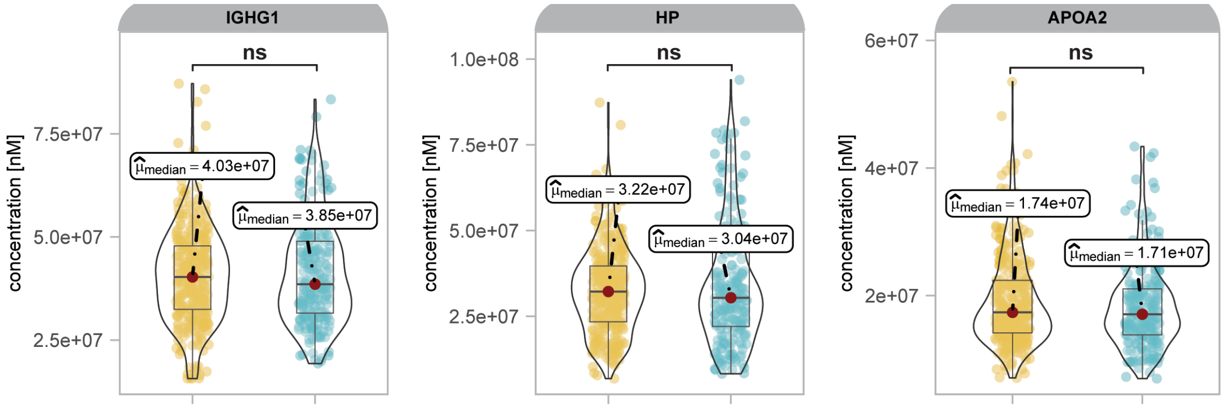


**Supplementary Figure 1. Plasma protein levels of three illustrative proteins (IGHG1, HP and APOA2) that do not show any differences in median protein concentrations between female and male donors across the ~600 measured samples.** The NS signifies that the proteins have no significant p-value (BH adjusted) between male and female donors, above 0.05.

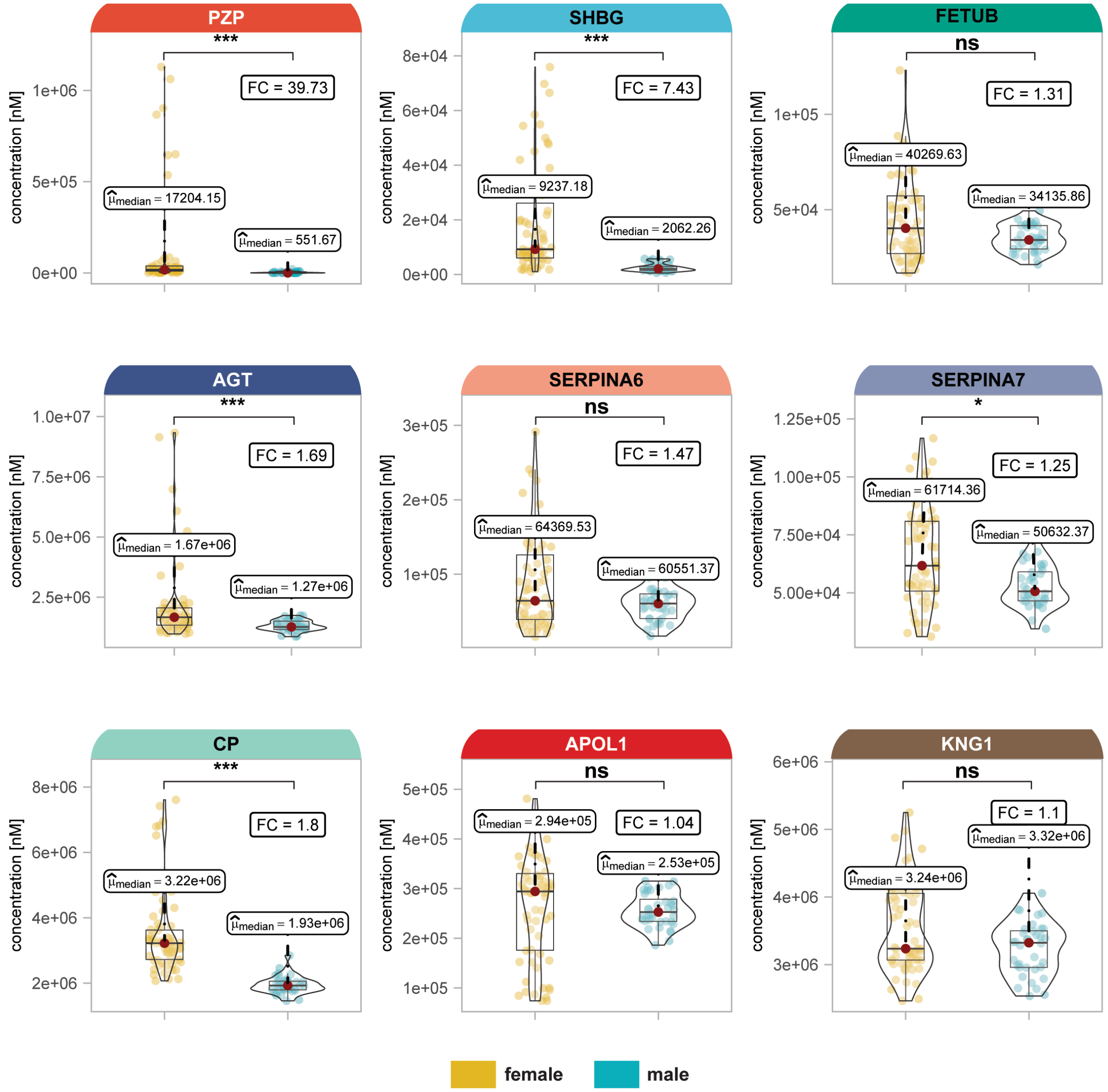


**Supplementary Figure 2. Differences between female and male plasma protein concentrations in the AICOVI cohort.** Median concentrations of 9 proteins in 11 participants sampled at 9 time points in the AICOVI study. One star (*) represents p-value (BH adjusted) below 0.05, two stars (**) below 0.01, and three stars (***) below 0.001, NS represents non-significant p-values above 0.05. In the TIMES cohort these proteins were significantly more abundant in the plasma proteome of women compared to men. Some of the proteins like FETUB, SERPINA6, APOL1, KNG1 do not differ significantly between women and men the AICOVI cohort, likely due to the smaller sample size.

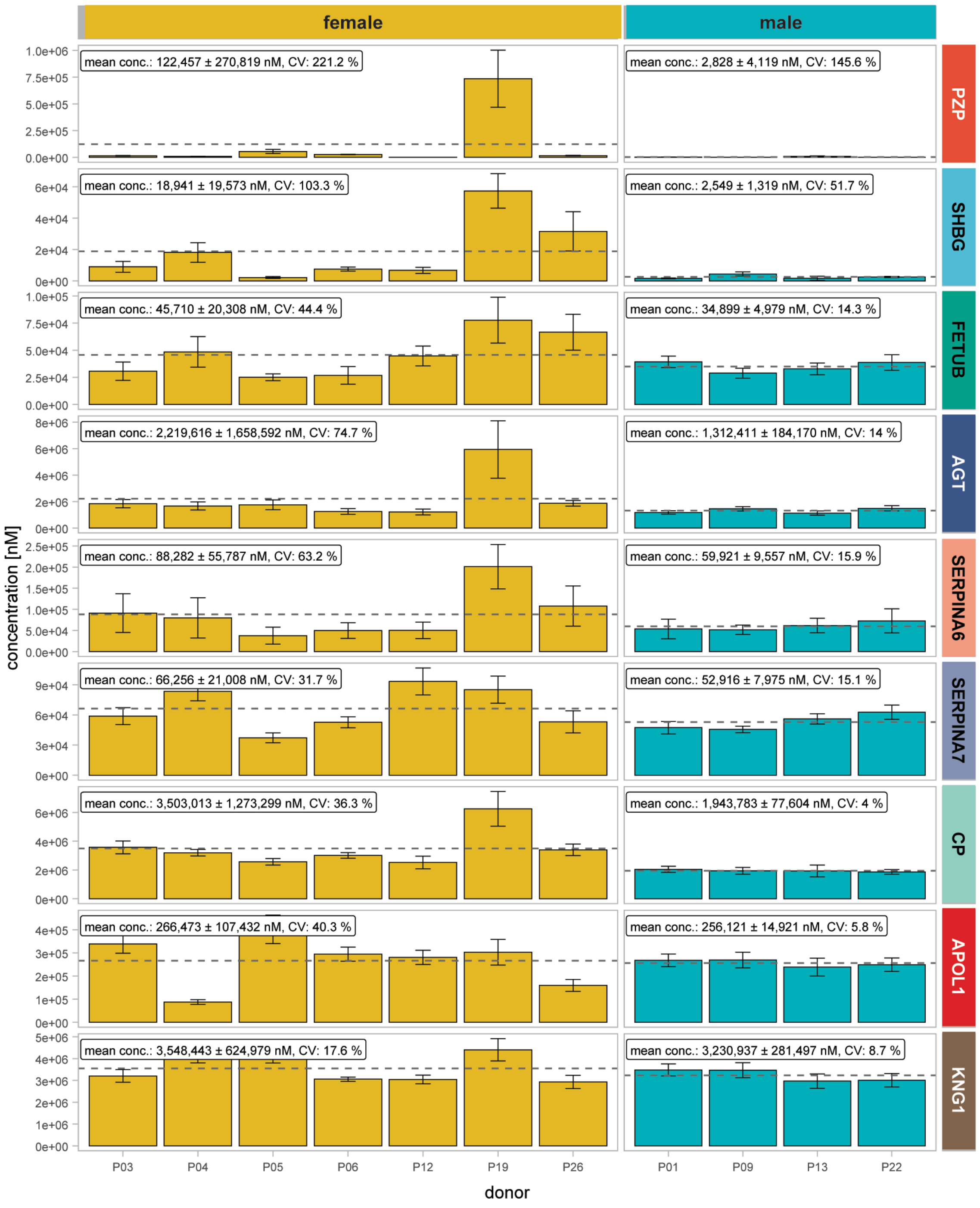


**Supplementary Figure 3. Mean plasma concentrations of nine selected proteins (PZP, SHBG, FETUB, AGT, SERPINA6, SERPINA7, CP, APOL1 and KNG1) in female and male donors in the AICOVI cohort.** The barplot with standard deviations depicts plasma concentrations of the nine selected proteins per female (yellow) and male (blue) donors within the AICOVI cohort. The dashed lines signify the overall mean concentrations in females and males, respectively. In each sub-figure the mean concentration is given at the top together with the coefficient of variation (CV).

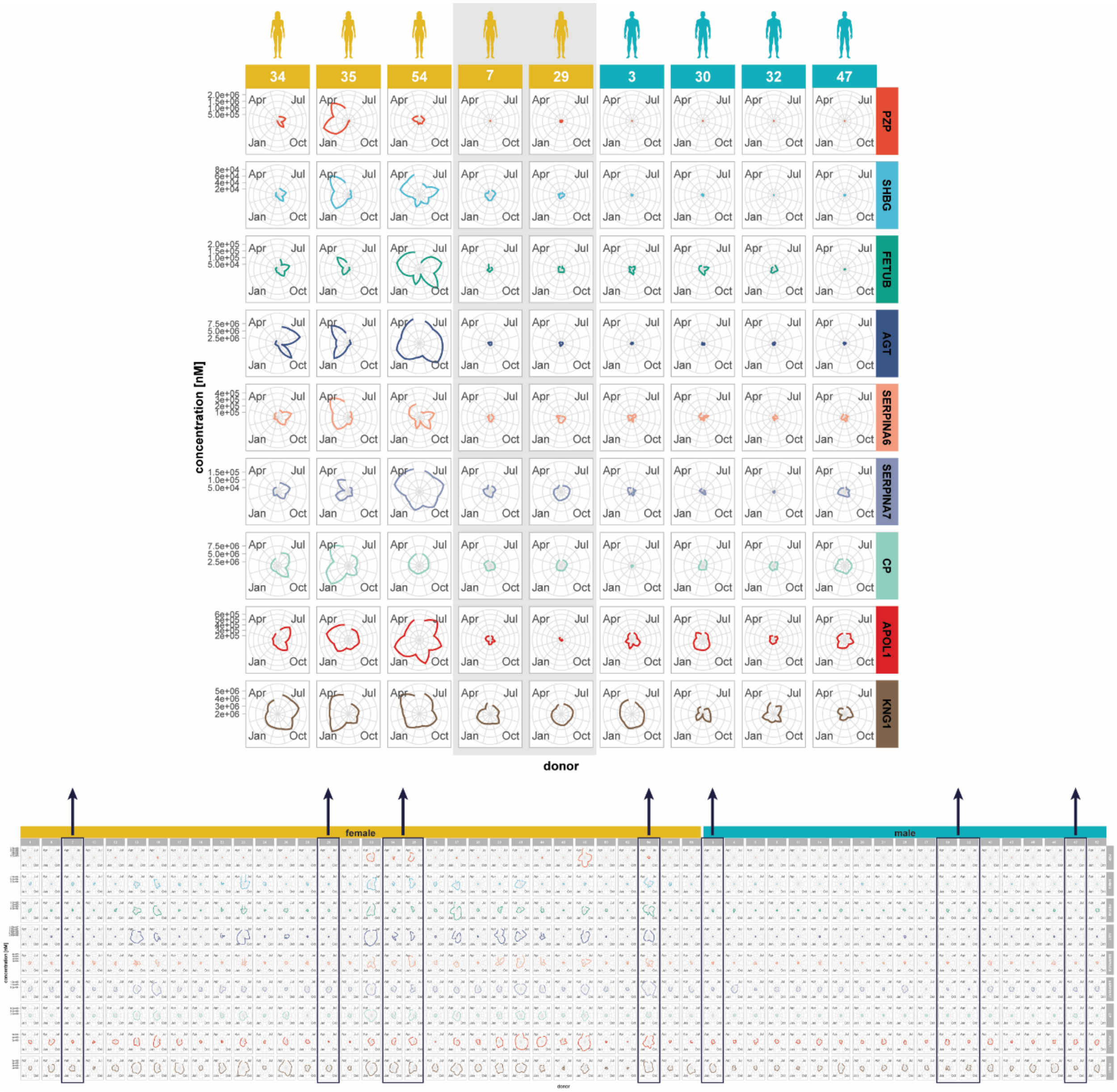


**Supplementary Figure 4. Radar plots of nine selected proteins (PZP, SHBG, FETUB, AGT, SERPINA6, SERPINA7, CP, APOL1 and KNG1) in all (panel below) and selected (panel above) donors.** Radar plots of selected proteins across all donors in TIMES cohort, divided into females (yellow) and males (blue) are shown in the lower panel. The upper panel represents the same donors that are also shown in the main Figure 3. These are boxed in the lower panel.

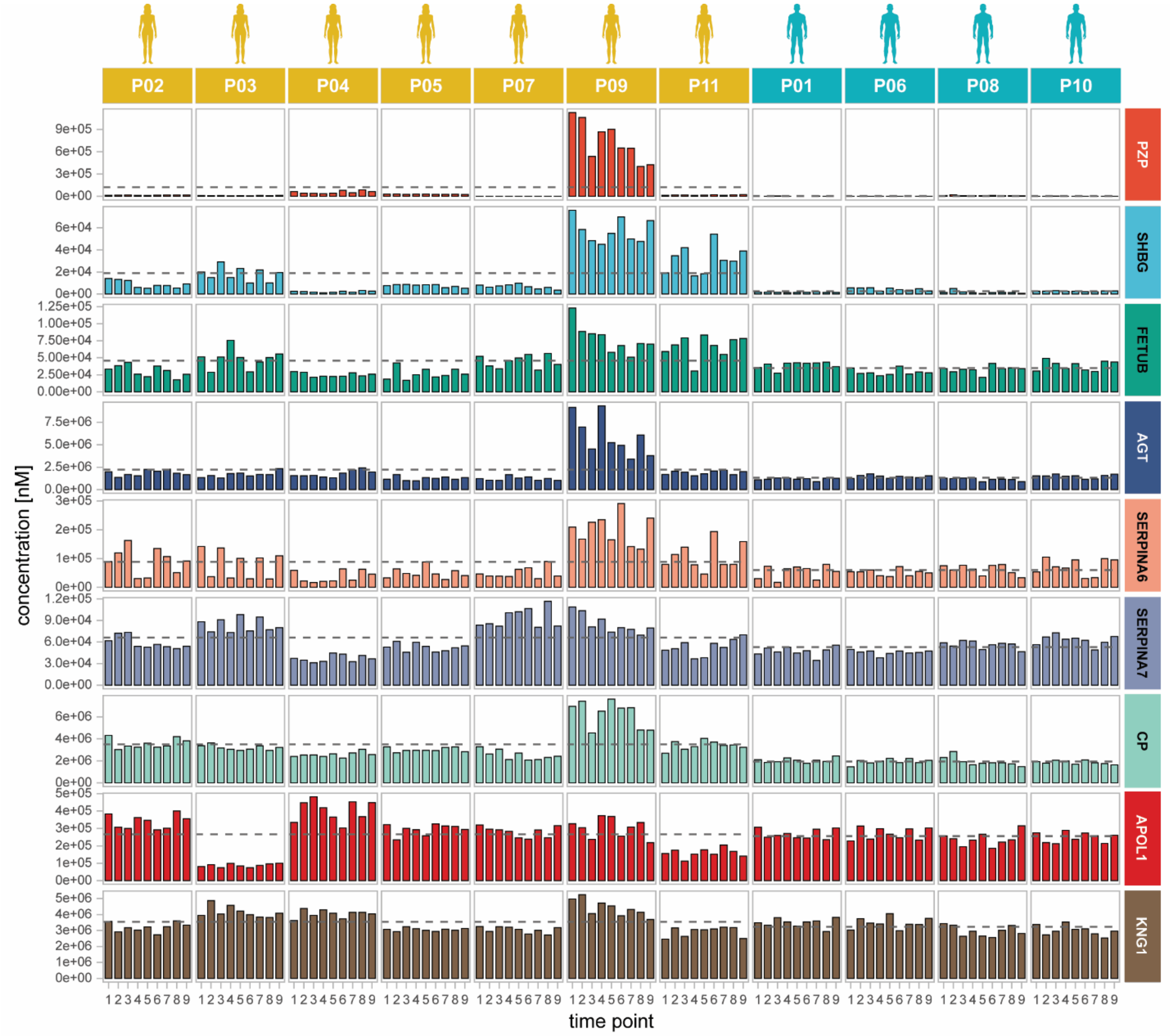


**Supplementary Figure 5. Bar plots of the concentrations of nine selected plasma proteins (PZP, SHBG, FETUB, AGT, SERPINA6, SERPINA7, CP, APOL1 and KNG1) in the 11 donors of the AICOVI study.** Bar plots of measured protein concentrations in the plasma of the 11 AICOVI donors reveal temporal bursts and declines. Each graph represents the concentration at each of the 9 time points of sampling.

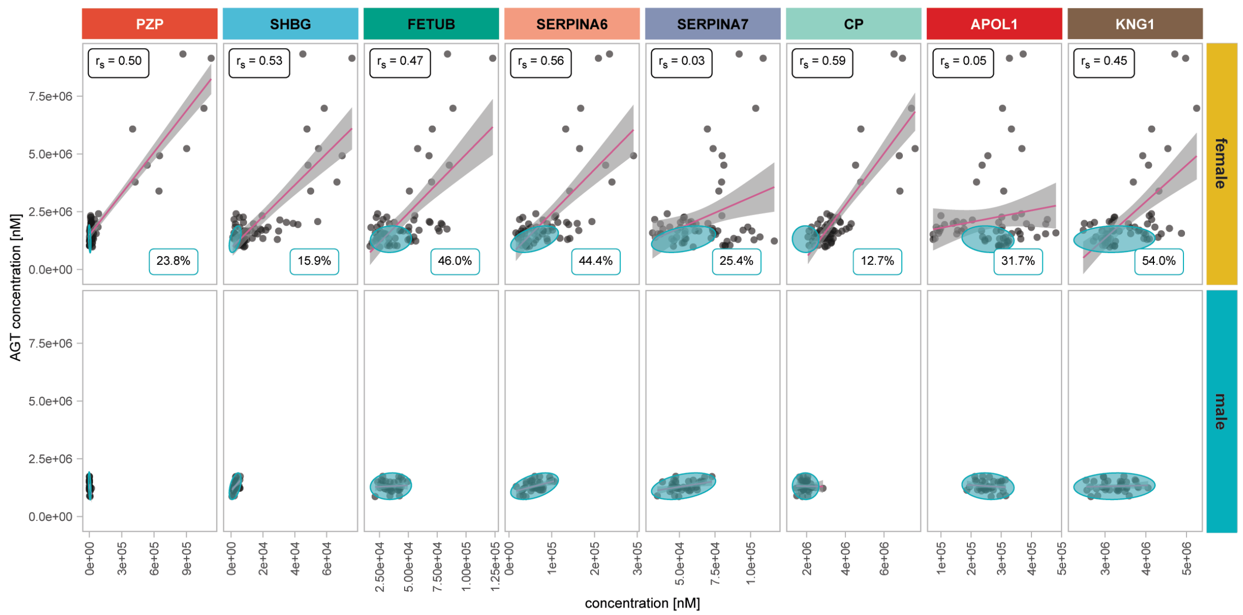


**Supplementary Figure 6. Correlation (r) plots of eight selected plasma proteins (PZP, SHBG, FETUB, SERPINA6, SERPINA7, CP, APOL1 and KNG1) with AGT in female and male individual donor samples of the AICOVI cohort.** The blue ellipses indicate the 95% confidence region of the male AGT-protein distribution, calculated assuming a bi-variate normal distribution. The percentage of female donor samples that fall into that ellipse is indicated in the bottom of the top panel.
